# Internal validation of models that estimate Food Intake LEVEL Scale thresholds after stroke using routinely available clinical variables

**DOI:** 10.1101/2024.03.11.24304069

**Authors:** Kazuhiro Okamoto, Keisuke Irie, Kengo Hoyano, Isao Matsushita

**Affiliations:** Department of Rehabilitation, Faculty of Health Science, Fukui Health Science University, Fukui, Japan; Cognitive Motor Neuroscience, Department of Human Health Sciences, Graduate School of Medicine, Kyoto University, Kyoto, Japan; Department of Rehabilitation Medicine, Kanazawa Medical University, Ishikawa, Japan

**Keywords:** Decision curve analysis, Deglutition disorders, Functional Independence Measure, Rehabilitation nursing, Swallowing

## Abstract

**Objectives:** To develop and internally validate models that estimate swallowing status after acute stroke by using routinely available clinical variables.

**Methods:** This retrospective observational study included patients with first-ever stroke who underwent a videofluoroscopic swallowing study or fiberoptic endoscopic evaluation of swallowing during hospitalization. Two binary outcomes were defined at the index assessment: Food Intake LEVEL Scale (FILS) ≥ 3 and FILS ≥ 7. The candidate predictors included age, sex, stroke type, Japan Coma Scale, modified Rankin Scale, Functional Independence Measure motor and cognition, serum albumin level, body mass index, and time from stroke onset. Performance was evaluated using the area under the receiver operating characteristic curve (AUC), calibration, and decision curve analysis (DCA). Stability selection was used to examine the predictor consistency.

**Results:** Among 126 patients, 112 (88.9%) and 73 (57.9%) patients had FILS ≥ 3 and FILS ≥ 7 at the index assessment, respectively. The out-of-fold AUCs were 0.759 and 0.821 for FILS ≥ 3 and FILS ≥ 7, respectively. DCA suggested greater clinical utility for the FILS ≥ 7 model than for the FILS ≥ 3 model. Stability selection showed that cognitive and functional measures were consistently important for FILS ≥ 3, whereas albumin and time from onset additionally contributed to FILS ≥ 7.

**Conclusions:** Routinely available clinical variables may help estimate swallowing status after acute stroke when dysphagia-specific assessments are unavailable or incomplete. The FILS ≥ 7 model showed the most favorable potential clinical utility and should be externally validated.

## 1. Introduction

Stroke remains a major cause of disability and mortality worldwide and often leads to complex clinical management in acute care, particularly among older adults.^1^ Dysphagia is a frequent complication after stroke and is associated with aspiration pneumonia and inadequate oral intake, contributing to malnutrition, prolonged hospitalization, and poorer recovery.^2,3^ Accordingly, the timely assessment of swallowing ability and the appropriate management of nutrition intake are important components of acute stroke care.

In numerous healthcare settings, particularly during off-hours such as weekends, access to dysphagia specialists is limited. Thus, nursing staff play an important role in preliminary dysphagia assessment. As primary caregivers, they can identify swallowing difficulties and initiate early interventions. Additionally, 24/7 dysphagia screening by proficiently trained nurses can reduce aspiration pneumonia incidence in hospitalized patients with stroke, reducing the duration of hospitalization.^4,5^ However, a substantial proportion of patients with acute stroke have reduced consciousness, cognitive impairment, or limited cooperation, and some are considered to have a high aspiration risk.^6,7^ In such situations, structured screening protocols and instrumental examinations (e.g., videofluoroscopic swallowing study [VFSS] and fiberoptic endoscopic evaluation of swallowing [FEES]) may not be feasible or may be deferred.^8^ Simultaneously, nurses are continuously present at the bedside and are expected to support early risk identification and management^9–11^ despite the increasing workload and staffing constraints reported in many healthcare systems. These realities highlight the need for practical approaches that support clinical decision making when swallowing evaluations cannot be completed in a timely manner.^12^

In this study, we intentionally focused on variables that are routinely collected or readily reviewed in the medical records of acute care settings rather than dysphagia-specific examination items. Specifically, we examined whether non-dysphagia-specific measures, such as functional status measures and general laboratory markers, could be used to estimate swallowing ability by nurses at the time of clinical assessment. Direct swallowing screening components or specialized physiological measures were not included because these tests require dedicated equipment, time, or specific training and are not consistently available or performed for all patients in routine practice. This study does not aim to replace comprehensive swallowing evaluation when available but addresses clinical situations where such evaluations are unavailable or incomplete.

Importantly, this study did not aim to predict whether a patient would eventually reach a given swallowing level. Instead, our goal was to estimate the swallowing ability at the time of clinical assessment by using information available in routine care. This distinction is important because the predictors and intended uses differ between estimating the current status and predicting later recovery. Thus, a model intended for contemporaneous estimation is needed as a clinical decision-support tool for current management and not as a prognostic instrument.

Several studies have developed prediction models for dysphagia, aspiration risk, or enteral feeding after stroke by using neurological severity, functional measures, and laboratory variables. Recent studies have increasingly reported more complete performance metrics, including calibration.^13,14^ Nevertheless, many published models rely on dysphagia-specific measurements or structured screening components that may be unavailable to all patients in routine acute care. Therefore, a practical gap remains in determining whether swallowing status can be estimated using only low-burden variables that are routinely available and assessed by nursing staff. To support clinical translation, prediction models should undergo rigorous internal validation and be reported with discrimination, calibration, and decision-analytic utility measures.^15–17^ Given the modest sample size of this study, we applied nested cross-validation and transparent reporting to reduce optimism and support credible internal validation.

We aimed to develop and internally validate models that estimate contemporaneous swallowing status in patients with stroke by using routinely collected clinical variables. We evaluated model performance by using nested cross-validation, reported discrimination and calibration, and examined decision-analytic utility by using decision curve analysis (DCA). Additionally, we performed stability selection to identify the variables that consistently contributed to the estimation across repeated subsamples. By focusing on information widely available to nursing staff and clinical teams, we seek to support practical decision making in acute stroke care when comprehensive swallowing evaluation is not feasible.

## 2. Materials and Methods

### 2.1. Study design

This retrospective observational study was conducted in accordance with the Strengthening the Reporting of Observational Studies in Epidemiology statement^18^ for observational studies and the Transparent Reporting of a multivariable prediction model for Individual Prognosis Or Diagnosis statement for the development and internal validation of multivariable prediction models.^19^ The primary objective was to estimate the contemporaneous swallowing status at the time of assessment rather than to predict subsequent recovery. We used routinely collected clinical data from patients with acute stroke admitted to Kanazawa Medical University Hospital between October 1, 2017, and October 31, 2020. The study protocol was approved by the Institutional Review Board (approval number: 78) and was conducted in accordance with the Declaration of Helsinki. The requirement for written informed consent was waived because of the retrospective nature of the study.

### 2.2. Patients

Eligible participants were adult patients with first-ever stroke who underwent VFSS or FEES as part of their routine clinical care during hospitalization. These examinations were used for clinical management; however, no dysphagia-specific test variables were included as predictors in the present models, which were developed using routinely available non-dysphagia-specific variables. We excluded patients with subarachnoid hemorrhage, brainstem or cerebellar lesions, prior head and neck malignancy or esophageal cancer treatment, pre-stroke permanent tube feeding, or missing data in the outcome or prespecified predictors. To ensure contemporaneous estimation, when multiple swallowing assessments were available for a single patient, we selected the first recorded assessment within 60 days of admission as the index assessment. Predictors were extracted from the same assessment episode or within a predefined time window anchored to the Food Intake LEVEL Scale (FILS)^20^ assessment (within 24–48 hours). Patient identification and exclusions as well as the final analytic cohort are summarized in the study flow diagram (Figure S1).

### 2.3. Outcome definition

On the basis of the VFSS and FEES results,^21^ an initial nutrition and rehabilitation plan was established as part of routine care (mean time from stroke onset to examination: 5.61; standard deviation: 3.84 days). Swallowing status was measured using the FILS and recorded on an ordinal 10-point scale from 1 (no oral intake) to 10 (normal diet) by an experienced speech-language pathologist. The index FILS assessment was defined as the first FILS recorded within 60 days of admission, and the predictors were extracted within a predefined time window anchored to the FILS assessment. We defined two binary outcomes representing clinically actionable thresholds: FILS ≥ 3, which is commonly used as an operational boundary for suitability to initiate direct swallowing training under clinical supervision, and FILS ≥ 7, which is commonly used as an operational boundary for full oral intake without concomitant alternative nutritional support. These outcomes were treated as clinical states at the time of index assessment; the models were intended to estimate swallowing status at the time of index assessment rather than to forecast subsequent recovery.

### 2.4. Candidate predictors and rationale

Candidate predictors were prespecified on the basis of their feasibility and consistency of availability in routine acute stroke care, with a particular focus on information that nursing staff can obtain or review without additional specialized procedures. We intentionally restricted predictors to non-dysphagia-specific variables that are routinely documented (e.g., functional status measures and general laboratory markers) because dysphagia-specific screening items and specialized physiological measures require dedicated equipment, time, or training and are not consistently performed for all patients, particularly in those with impaired consciousness, cognitive dysfunction, limited cooperation, or clinical instability. Therefore, this study was designed for settings in which dysphagia-specific testing is unavailable or incomplete and does not intend to replace VFSS/FEES or comprehensive swallowing physiology testing when such evaluations are feasible. The prespecified predictors included age, sex, stroke type (ischemic or hemorrhagic), the Japan Coma Scale (JCS), the modified Rankin Scale (mRS), motor and cognitive subscores of the Functional Independence Measure (FIM), serum albumin (Alb), body mass index (BMI), and number of days from stroke onset to index assessment (time from onset). These variables were selected as candidate predictors on the basis of previous studies that reported associations with swallowing-related outcomes after stroke.^22–27^ The JCS was categorized into three levels (I, II, and III) to reflect clinically meaningful consciousness. Specifically, JCS 0–3 was coded as category I, JCS 10–30 as category II, and JCS 100–300 as category III. The categorical predictors were one-hot encoded. Continuous predictors were standardized using training data only within each cross-validation fold and then applied to the corresponding test data. Given that the FIM subscores and mRS both reflect functional status and may be correlated, we used regularized logistic regression and nested cross-validation to mitigate overfitting and reduce the sensitivity to collinearity.

### 2.5. Missing data

For each variable, we summarized missingness overall and stratified the results by outcome status for FILS ≥ 3 and FILS ≥ 7. For the primary analyses, missing numerical values were imputed using the median, and missing categorical values were imputed using the most frequent category. Imputation parameters were estimated using training data only within each cross-validation fold and then applied to the corresponding test data. We conducted sensitivity analyses by using complete-case data (excluding records with missing values in any candidate predictor used) and repeated model development without class weighting to assess the effect of class imbalance handling on the probability scale and calibration.

### 2.6. Model development for binary threshold models

We developed two separate prediction models for contemporaneous classification of FILS ≥ 3 and FILS ≥ 7. A regularized logistic regression with an elastic net penalty (SAGA solver) was used. This approach was selected to mitigate overfitting and handle collinearity among the predictors while retaining interpretability, which is appropriate for clinical use. We prespecified an interaction term between Alb and time from onset to assessment (Alb × time). This term represents a clinically plausible effect modification, whereby the association between a general nutritional marker and contemporaneous swallowing status may differ according to the timing after stroke onset.^28^ Alb and time from onset were mean-centered within the training folds before constructing the interaction to improve numerical stability and reduce collinearity with the main effects.

### 2.7. Internal validation and performance estimation

Considering that model development with modest sample sizes is vulnerable to overfitting and optimistic performance estimates, we used nested cross-validation to separate hyperparameter selection from performance estimation. In the outer loop, we performed repeated stratified 5fold cross-validation (10 repeats) to obtain the out-of-fold predicted probabilities for each participant. In the inner loop, we performed a stratified five-fold cross-validation for hyperparameter tuning by using grid search. The regularization strength and elastic net mixing parameters were tuned to maximize the area under the receiver operating characteristic (ROC) curve (AUC) of the inner folds. We pooled out-of-fold predictions from the outer loop to estimate the overall performance. Discrimination was quantified using the AUC. The Brier score was reported as an overall measure of the accuracy of the predicted probabilities.

### 2.8. Probability calibration and decision-analytic utility

Given that the predicted probabilities were intended to support clinical decision making, we evaluated calibration by using reliability diagrams (calibration plots) based on out-of-fold predictions. Calibration was performed using training data only within each outer-fold training set by cross-validated sigmoid calibration (Platt scaling), and isotonic calibration was considered only when the number of events supported stable estimation. The calibrated model was then applied to the corresponding outer-fold test data. We evaluated clinical usefulness by using DCA based on out-of-fold predicted probabilities. Net benefit was calculated over a prespecified range of threshold probabilities (0.05–0.95) and was compared with the results of the “treat all” and “treat none” strategies. The interpretation focused on the threshold probability ranges that are plausible for acute stroke decision making, where the threshold probability represents the point at which clinicians would choose to act on the basis of the predicted probability. Although we present net benefit across a wide range (0.05–0.95), interpretation was centered on clinically plausible thresholds where swallowing management and nutritional strategy decisions are typically considered in acute stroke care.

### 2.9. Stability selection

To identify the predictors that consistently contributed to the estimation across the resampled datasets, we performed stability selection by using L1-regularized logistic regression, specifically least absolute shrinkage and selection operator. For each outcome (FILS ≥ 3 and FILS ≥ 7), we generated 500 random subsamples without replacement (subsample fraction=0.75). In each subsample, we refit the model by using the same preprocessing steps (including one-hot encoding and a prespecified interaction term). For each transformed feature, we calculated the selection frequency, which was defined as the proportion of subsamples in which the coefficient was non-zero. We reported the highest-frequency features separately for the FILS ≥ 3 and FILS ≥ 7 models.

### 2.10. Supplementary ordinal analysis

We examined the ordinal-level agreement across the 10-level FILS by using an internally validated ordinal surrogate approach. We modeled FILS as a continuous outcome by using a gradient boosting regression model with absolute error loss and then rounded and clipped the predicted values to the 1–10 range to obtain ordinal estimates. The performance was evaluated using out-of-fold predictions (i.e., predictions for participants not used to fit the model in that fold), summarized by the mean absolute error (MAE; in grades) and quadratic weighted kappa (QWK), which penalizes larger disagreements more heavily. Agreement was visualized using a confusion matrix displayed as a heatmap. Given that the primary aim of this study was threshold-based decision support, ordinal analysis was supplementary and implemented as a pragmatic internally validated surrogate approach rather than as a primary ordinal regression model.

All analyses were performed using Python 3.10 (Python Software Foundation, Wilmington, Delaware, USA) with scikit-learn and related scientific libraries. All preprocessing steps, including imputation, scaling, encoding, interaction generation, and calibration, were fitted using training data only within each cross-validation fold and then applied to the corresponding test data.

## 3. Results

### 3.1. Study sample and outcome distribution

Table 1 summarizes the patient details and predictive variables. For the prespecified clinically actionable thresholds, 112/126 patients (88.9%) and 73/126 patients (57.9%) had FILS ≥ 3 and FILS ≥ 7 at the index assessment, respectively.

**Table 1.**
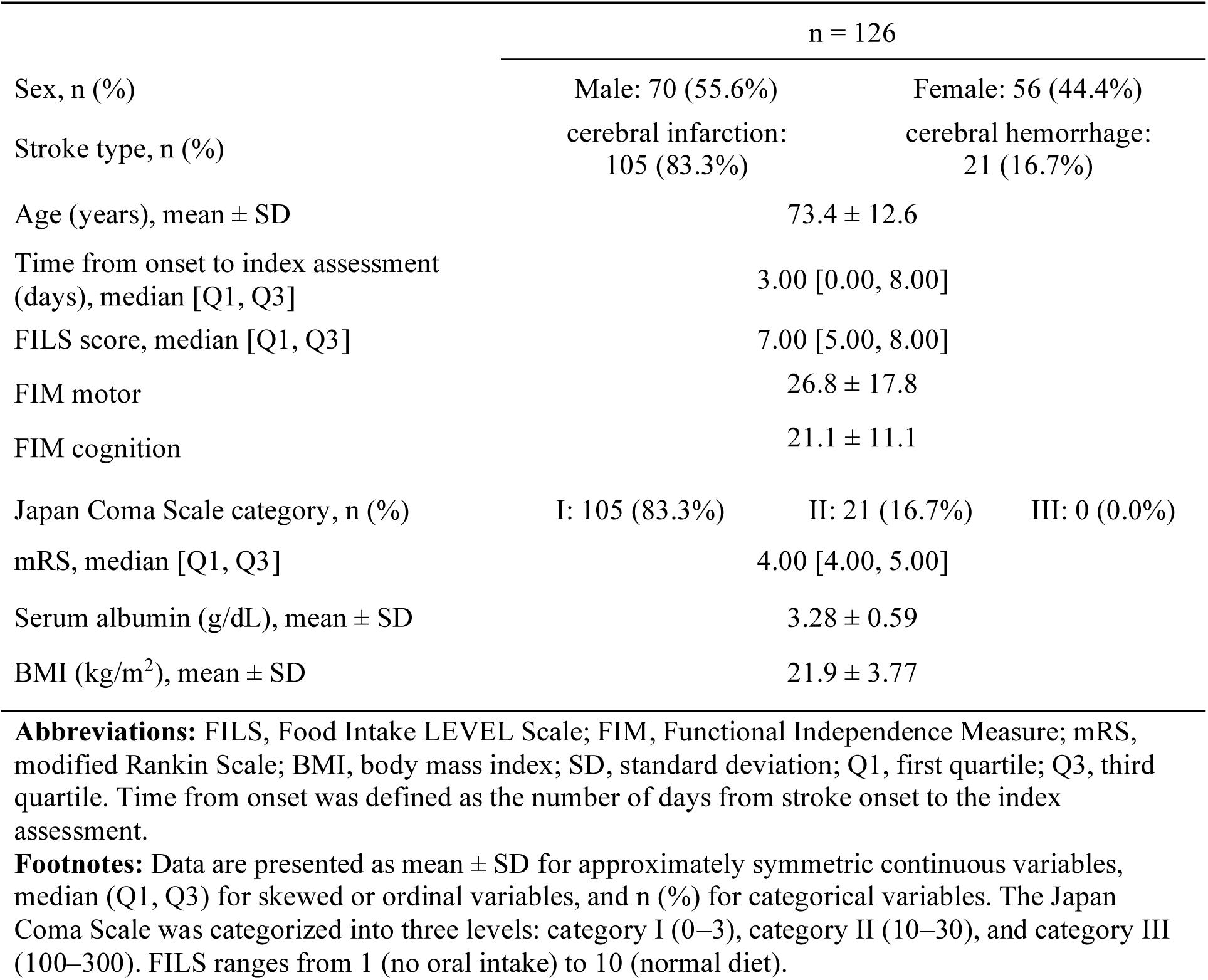
Baseline characteristics of the study participants.

### 3.2. Missing data

Among the prespecified candidate predictors used in the main models (age, sex, stroke type, JCS, mRS, FIM motor and cognition, Alb, BMI, and time from onset), there were no missing values (0% missingness for each variable). Accordingly, the primary analyses were not materially influenced by imputation, and the complete-case sensitivity analyses yielded the same analytical sample size.

### 3.3. Sensitivity analyses

Table S1 lists the results of the sensitivity analysis. Given that there were no missing values in the prespecified predictors, complete-case analyses yielded performance estimates identical to those of the primary analysis. Removing the class weighting produced similar discrimination and Brier scores for both thresholds.

### 3.4. Discrimination of the threshold models

By using nested cross-validation, the model for FILS ≥ 3 achieved an out-of-fold AUC of 0.759, indicating moderate discrimination. The model for FILS ≥ 7 achieved an out-of-fold AUC of 0.821, indicating good discrimination. Figure 1 shows the ROC curves for both threshold models.

**Figure 1.**
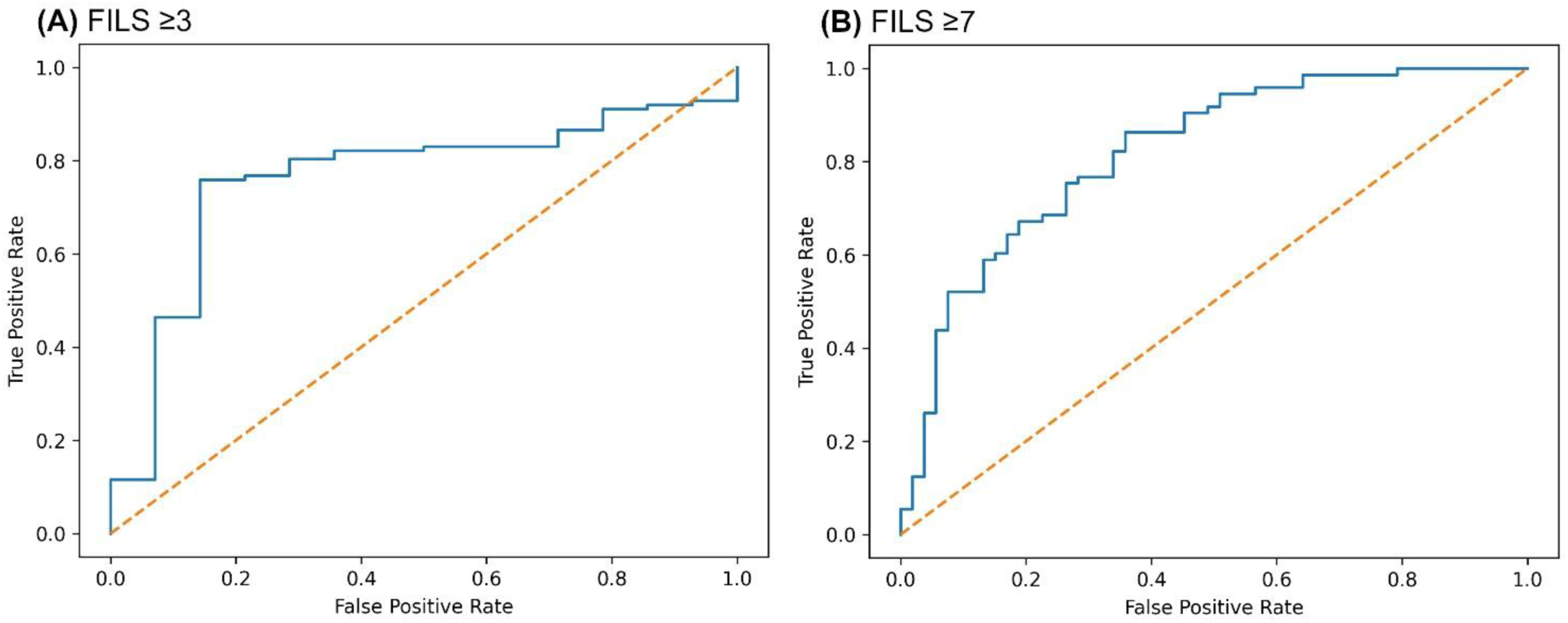
ROC curves for the threshold models based on out-of-fold predictions from nested cross-validation. **(A)** FILS ≥ 3. **(B)** FILS ≥ 7. AUC values are reported in Table 2. ROC, receiver operating characteristic; FILS, Food Intake LEVEL Scale; AUC, area under the receiver operating characteristic curve

**Table 2.**
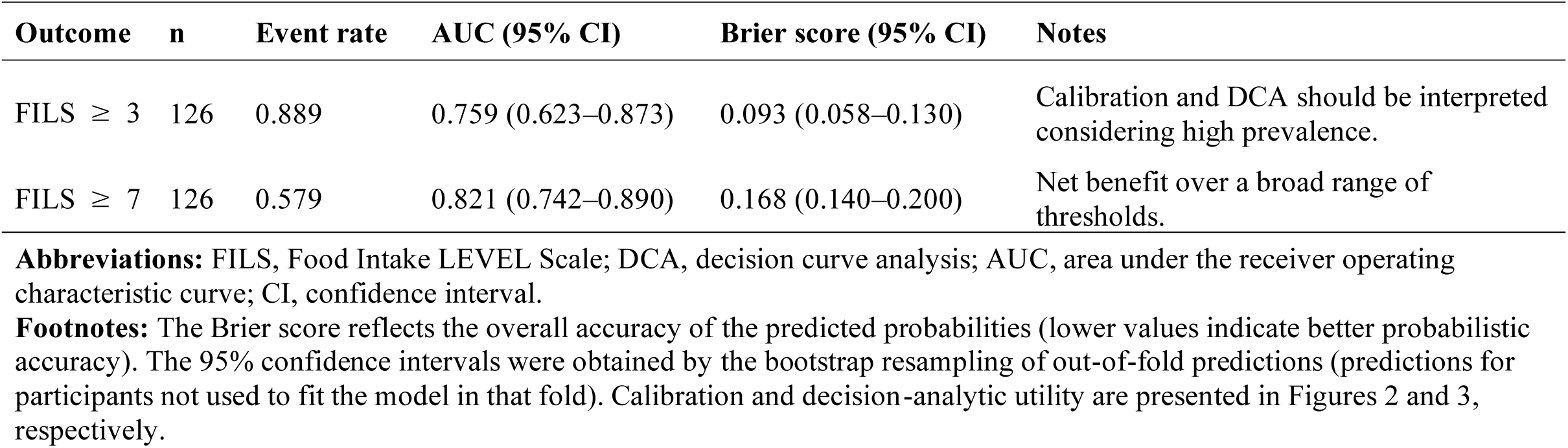
Model performance for estimating contemporaneous swallowing status.

Table 2 summarizes the overall model performance. In addition to discrimination, probabilistic accuracy was assessed using the Brier score, which was 0.093 (95% CI 0.058–0.130) for FILS ≥ 3 and 0.168 (95% confidence interval [CI]=0.140–0.200) for FILS ≥ 7 according to out-of-fold predictions from nested cross-validation.

### 3.5. Calibration and decision-analytic utility

Calibration was evaluated using calibration plots based on out-of-fold predicted probabilities (Figure 2). FILS ≥ 3 interpretation should be cautious because outcome prevalence was high (88.9%), limiting the stability of probability estimates at the extremes. DCA was performed to assess the potential clinical usefulness across a range of threshold probabilities (Figure 3).

**Figure 2.**
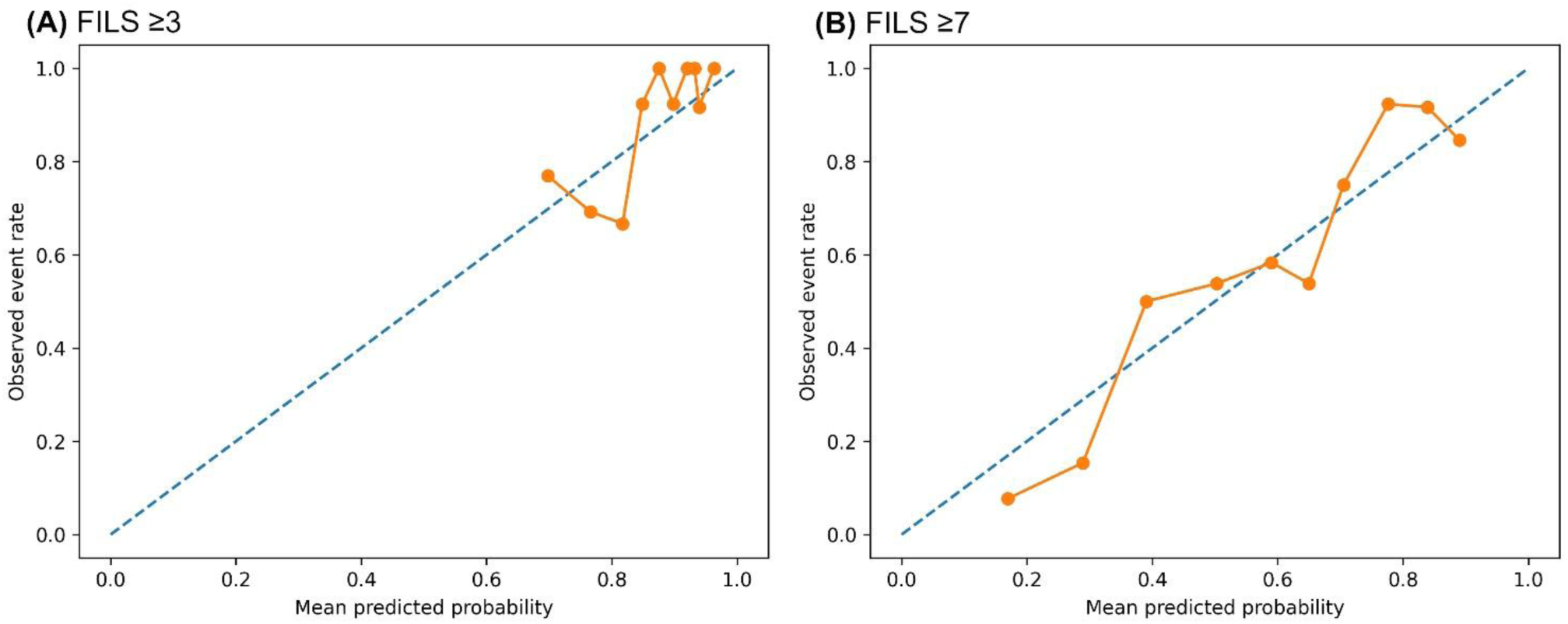
Calibration plots for the threshold models based on out-of-fold predicted probabilities from nested cross-validation. **(A)** FILS ≥ 3. **(B)** FILS ≥ 7. The diagonal line indicates perfect calibration; points represent observed event proportions within bins of predicted probability. FILS, Food Intake LEVEL Scale

**Figure 3.**
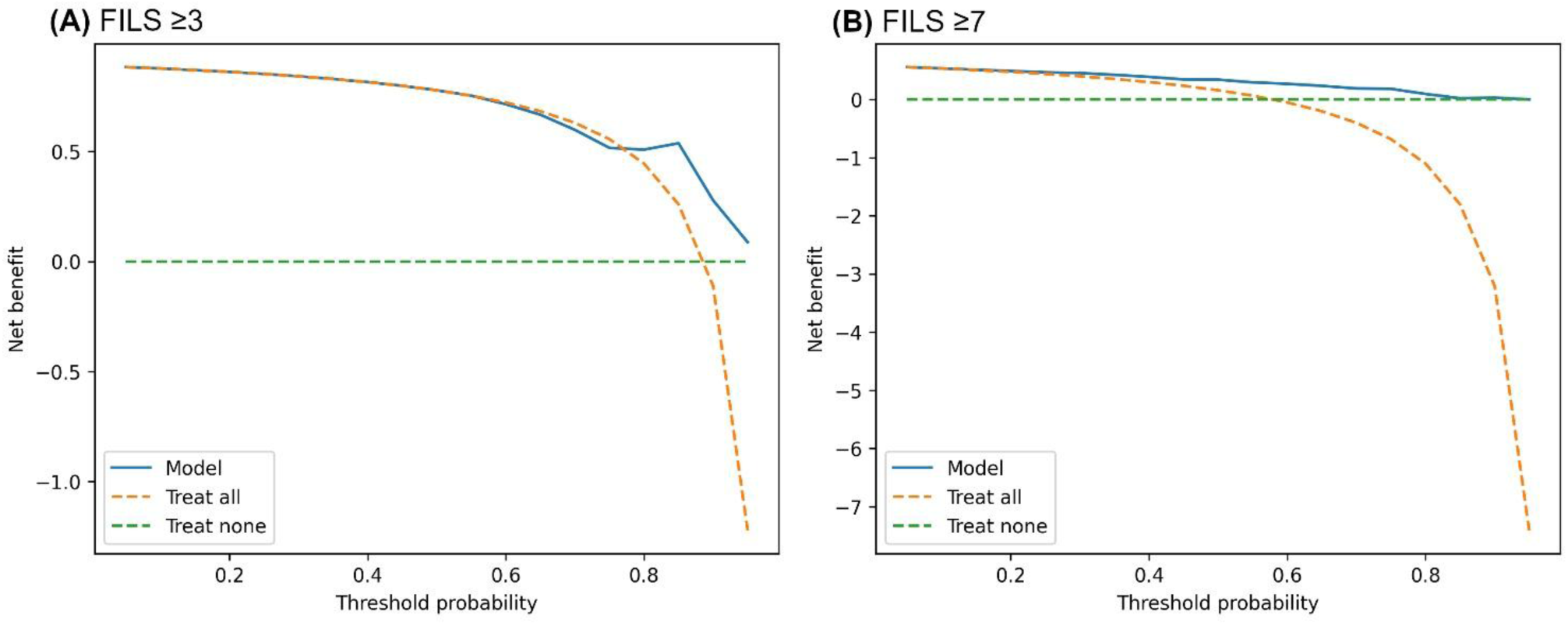
DCA for the threshold models based on out-of-fold predicted probabilities from nested cross-validation. **(A)** FILS ≥ 3. **(B)** FILS ≥ 7. The x-axis shows the threshold probability (the probability at which a clinician would choose to act), and the y-axis shows net benefit. Curves are compared with “treat all” and “treat none” strategies. Note that the y-axis scale differs between panels to improve readability because the “treat all” strategy yields large negative net benefit at high threshold probabilities. DCA, decision curve analysis; FILS, Food Intake LEVEL Scale

For FILS ≥ 7, the model showed net benefit compared with both “treat all” and “treat none” strategies across a broad range of threshold probabilities, thus supporting potential utility when clinical decisions are guided by a probability-based rule. For FILS ≥ 3, where outcome prevalence was high, the “treat all” strategy performed strongly at lower threshold probabilities; contrastingly, the model’s incremental net benefit was more evident at higher threshold probabilities, where avoiding unnecessary action becomes more important.

### 3.6. Stability selection

Stability selection (500 subsamples; subsample fraction: 0.75) was used to identify the predictors that consistently contributed to each threshold model (Table 3).

**Table 3.**
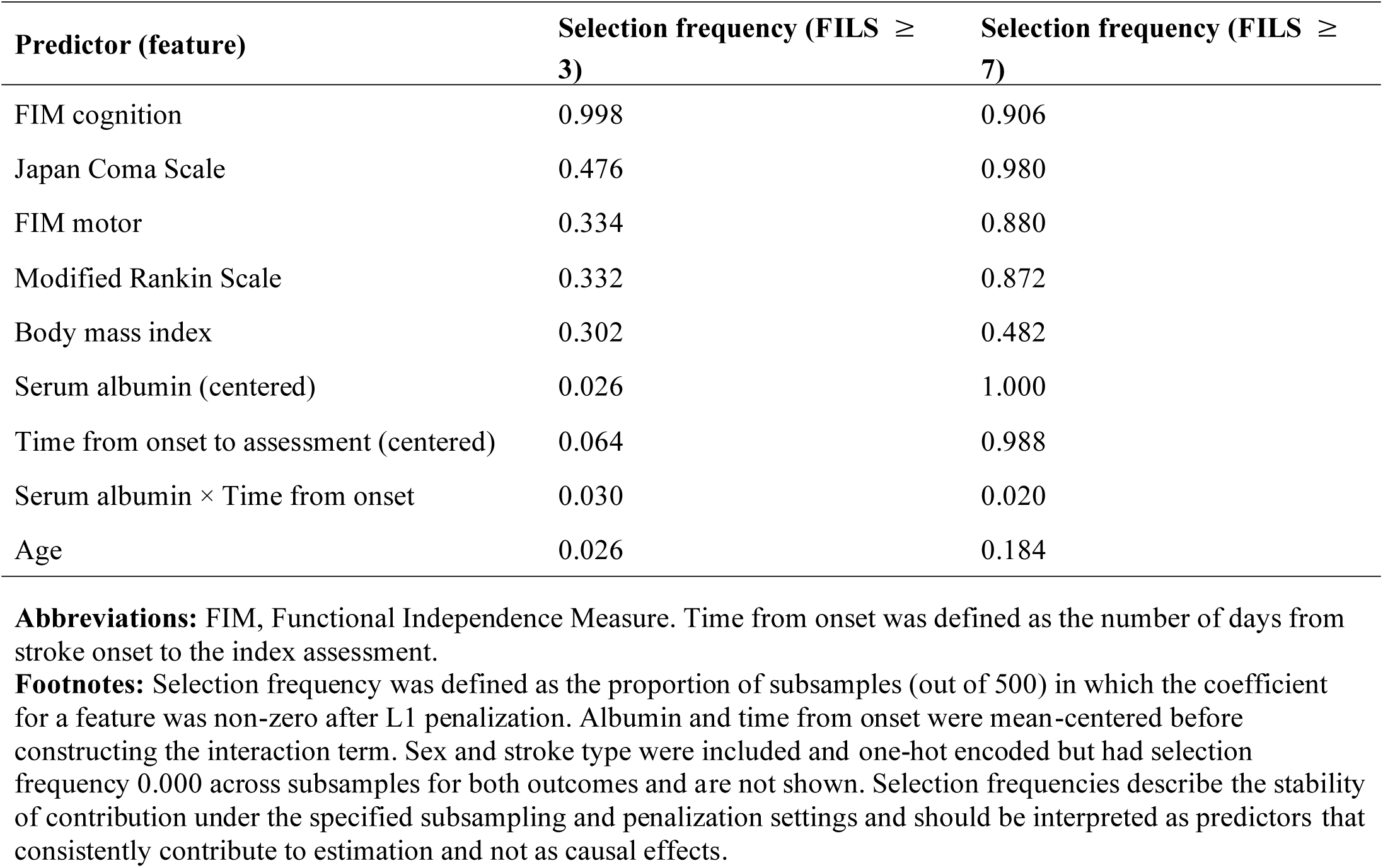
Stability selection results for the threshold models.

For FILS ≥ 3, FIM cognition was selected in nearly all subsamples (selection frequency: 0.998). JCS (0.476), FIM motor (0.334), mRS (0.332), and BMI (0.302) were selected with moderate frequency. Time from onset (0.064), Alb (0.026), age (0.026), and Alb × time (0.030) were infrequently selected. Sex and stroke type were not selected (0.000).

For FILS ≥ 7, Alb was selected in all subsamples (1.000). Time from onset (0.988), JCS (0.980), FIM cognition (0.906), FIM motor (0.880), and mRS (0.872) were selected with high frequency. BMI showed moderate selection frequency (0.482), age showed low selection frequency (0.184), and the interaction term Alb × time was infrequently selected (0.020). Sex and stroke type were not selected (0.000).

Overall, the stability selection profiles differed according to the threshold. FILS ≥ 3 was characterized primarily by cognitive, alertness-related, and functional measures, whereas FILS ≥ 7 additionally showed strong and consistent contributions from serum Alb and duration since stroke onset.

### 3.7. Supplementary ordinal analysis

Ordinal-level agreement across the 10-level FILS was assessed using an internally validated surrogate ordinal approach. The out-of-fold MAE was 1.437, and the QWK was 0.622, thus indicating moderate agreement between the predicted and observed FILS categories. The agreement was visualized using a confusion matrix displayed as a heat map (Figure S2).

## 4. Discussion

In this retrospective study, we developed and internally validated prediction models to estimate contemporaneous swallowing status after acute stroke by using routinely available, non-dysphagia-specific clinical variables. We focused on FILS ≥ 3 and FILS ≥ 7 at the index assessment. Discrimination was moderate for FILS ≥ 3 (AUC=0.759) and good for FILS ≥ 7 (AUC=0.821). DCA suggested that the FILS ≥ 7 model provided a net benefit compared to the “treat all” and “treat none” strategies across a broad range of threshold probabilities. Contrastingly, for FILS ≥ 3, the incremental advantage compared with a “treat all” approach was less pronounced at lower thresholds; this finding is consistent with the high prevalence of FILS ≥ 3 in this cohort. This study was designed for routine acute care settings. In many acute stroke settings, instrumental examinations such as VFSS/FEES and structured dysphagia screening may be delayed or incomplete because of off-hour staffing constraints, limited specialist availability, clinical instability, impaired consciousness, cognitive dysfunction, or limited cooperation. Accordingly, clinical teams still need to approximate swallowing status to support timely rehabilitation planning and nutritional strategies while recognizing that any estimation tool is complementary to, rather than a replacement for, comprehensive swallowing evaluation. Therefore, we intentionally restricted the candidate predictors to variables that are routinely documented and can be obtained or reviewed with minimal additional burden. This design addresses a recurring practical gap in acute care: dysphagia-specific measures may be informative when available, but they are not consistently obtained for all patients, particularly those who cannot safely participate in swallowing tasks or adhere to testing protocols.^29^ The stability selection results provided clinically interpretable insights into how the predictor profiles differed between the two thresholds. For FILS ≥ 3, FIM cognition was selected in nearly all subsamples (selection frequency 0.998), whereas JCS, FIM motor, mRS, and BMI were selected with moderate frequency. By contrast, Alb levels and the duration since stroke onset were infrequently selected. This pattern aligns with the pragmatic clinical context of the FILS ≥ 3 threshold: decisions about initiating direct swallowing training may depend heavily on whether the patient can participate reliably and safely in therapy and follow instructions, which is closely linked to cognition and arousal. The prominence of FIM cognition also reflects the fact that measures of functional status capture the consequences of neurological impairment in a manner that is directly observable in daily care. Prior studies have supported the association between functional independence and swallowing-related outcomes in stroke and older populations, including the use of FIM as a prognostic tool and links between reduced activities of daily living and dysphagia.^30^ From an implementation standpoint, FIM has practical advantages in acute care because it is based on activities performed in everyday settings; furthermore, it can be assessed even when patients have impaired consciousness or cognitive dysfunction. Prior work has reported the acceptable reliability of FIM scoring when performed by non-specialist evaluators,^31,32^ thus supporting its suitability as a predictor in models designed to be feasible under routine clinical conditions. Although the current study was not designed to establish causal relationships, the consistency of FIM cognition across subsamples supports its relevance for estimating whether patients are likely to be above the lower functional threshold represented by FILS ≥ 3. This association may reflect the greater likelihood that patients with preserved cognitive function will participate more effectively in the assessment and rehabilitation of dysphagia.^33^

For FILS ≥ 7, the stability profile was distinct. Serum Alb level and duration since stroke onset were selected with very high frequency (1.000 and 0.988, respectively), in addition to the JCS and functional measures (FIM motor/cognition and mRS). This pattern suggests that estimating whether a patient is above the higher functional threshold (oral intake without alternative nutritional support) draws not only on the ability to participate in assessment, training, and alertness but also on the broader features of overall physiological status and early clinical course. Importantly, serum Alb levels should not be interpreted as a direct or specific marker of nutritional intake. Alb is influenced by multiple factors, including inflammation, illness severity, fluid balance, hepatic function, and acute-phase responses, and its relatively long half-life limits its sensitivity to short-term nutritional change.^34^ Therefore, in the present study, Alb was best viewed as an accessible, routinely recorded indicator of general systemic conditions that may correlate with the ability to sustain oral intake in an acute stroke setting. The duration since stroke onset may similarly reflect the evolving clinical trajectory during the early post-stroke period, including the stabilization of medical status and the accumulation of routine supportive care and rehabilitation exposure.^35^ Considering that the models were designed for contemporaneous estimation using low-burden variables rather than causal inference, these findings should be interpreted as practical associations useful for bedside estimation, not evidence that modifying Alb itself would change swallowing status.^36^ The FIM and mRS capture related but not identical aspects of functional status (domain-specific versus global disability), and regularization was used to reduce instability due to collinearity. Recent studies have shown that comprehensive physical exercise therapy for patients with stroke improves FIM motor scores and swallowing function.^37^ A study on older adults residing in long-term care facilities^30^ further supports this association by highlighting the relationship between reduced functional independence and dysphagia. Reduced functional independence is often correlated with stroke severity, and dysphagia may worsen because of not only impaired muscle output but also impaired pharyngolaryngeal sensation,^38^ poor posture,^39^ and reduced cough strength.^40^ Thus, the FIM and mRS, which reflect overall functional capacity, may serve as important indicators for predicting swallowing function.

DCA evaluates whether using predicted probabilities to guide an action would provide more benefits than simple default strategies across different probability thresholds.^41^ The threshold probability represents the point at which a clinician chooses to act on the basis of the model output. For FILS ≥ 3, the high prevalence of the outcome in our cohort means that “treat all” strategies can appear favorable at lower threshold probabilities, which is expected when most patients are classified as positive. Under such conditions, the added value of a model is more apparent in threshold regions where clinicians require higher certainty before acting and where reducing unnecessary actions matters more. For FILS ≥ 7, where prevalence was more balanced, the model showed a net benefit across a wide range of threshold probabilities compared with both “treat all” and “treat none,” thus supporting potential usefulness when clinicians must weigh the trade-off between advancing oral intake prematurely and unnecessarily continuing alternative nutrition. These findings emphasize that the anticipated value of a prediction model depends on the clinical decision context and the threshold at which clinicians would act rather than on discrimination metrics alone.

We prespecified the interaction term Alb × time as a clinically plausible modifier, but it was infrequently selected in the stability analyses for both thresholds. One possible explanation is that the main effects of Alb and time captured most of the information available in this dataset, thus leaving few stable incremental values for a linear interaction term under penalized modeling. Additionally, effect modification may not be well-represented by a single multiplicative term, and nonlinear relationships or larger samples may be required to reliably estimate such patterns. Importantly, the stability results do not support strong conclusions about the presence or absence of effect modification but suggest that within this dataset and modeling framework, the interaction did not contribute consistently to the estimation beyond the main effects.

This study has a few limitations that warrant consideration. First, this was a single-center retrospective study. The lack of external validation and reliance on data from a single center also raises concerns about the generalizability of the findings. Employing cross-validation minimizes analytical errors but does not compensate for the potential biases of a single-center study. Therefore, future studies should validate these models in multi-center studies by using published codes to ensure their reproducibility and reliability across different clinical settings. Second, the analytic cohort consisted of patients who underwent VFSS/FEES for clinical management. Although the models did not use dysphagia-specific test variables, selection bias may limit generalizability to patients who never underwent instrumental examination. Third, although the models were designed for contemporaneous estimation, the timing of predictor measurement relative to the index FILS assessment and local care pathways (including rehabilitation exposure and nutritional management) may influence model behavior, thus possibly reducing portability across institutions with different workflows. Fourth, the predictor set was intentionally restricted to routinely available non-dysphagia-specific variables. This supports the feasibility and addresses the specific implementation gap motivating this work. However, it also indicates that the models do not incorporate potentially informative dysphagia-specific measurements when available. Incorporating parameters such as tongue pressure,^42^ cough reflex strength, and practical swallowing screening test results^43–45^ may substantially improve predictive accuracy of the model. Future studies could evaluate a tiered approach in which the present low-burden model serves as a baseline and in which specialized dysphagia measures are added when feasible. Furthermore, studies should evaluate whether such augmentation improves performance and clinical utility without compromising applicability.

In conclusion, we developed internally validated models to estimate contemporaneous swallowing status after acute stroke by using routinely available clinical variables that nursing staff and clinical teams can readily obtain or review. The models demonstrated moderate-to-good discrimination, with more favorable decision-analytic utility for the higher threshold (FILS ≥ 7). Stability selection suggested distinct predictor profiles for FILS ≥ 3 and FILS ≥ 7, reflecting differences in the clinical context of these thresholds. These findings suggest that swallowing status can be estimated without dysphagia-specific testing, but require external validation in broader clinical populations.

## Acknowledgments

We extend our profound gratitude to Kaori Kyoda and Noriko Yamazaki of Kanazawa Medical University Hospital for their invaluable assistance in the measurement and compilation of patient data. We also express our sincere appreciation to the staff of Kanazawa Medical University Hospital. This study received no specific grants from any funding agency in the public, commercial, or not-for-profit sectors.

## Data availability

The analysis code and parameter settings are publicly available at https://github.com/kokamoto46/speech_audiology_prediction. The patient-level data used in this study are not publicly available because of ethical and privacy restrictions. De-identified data may be made available from the corresponding author upon reasonable request and with the approval of the Institutional Review Board of Kanazawa Medical University Hospital.

