## Supplemental Figures and Tables for "Internal validation of models that estimate Food Intake LEVEL Scale thresholds after stroke using routinely available clinical variables"

**Supplementary Figures and Tables**

This file contains two supplementary figures and one supplementary table that accompany the manuscript.

**Figure S1**, flow diagram of patient identification, exclusions, and the final analytic cohort;
**Figure S2**, confusion matrix (heatmap) for observed versus predicted FILS categories from the supplementary ordinal analysis;

**Table S1**, sensitivity analyses (nested cross-validation; out-of-fold predictions).

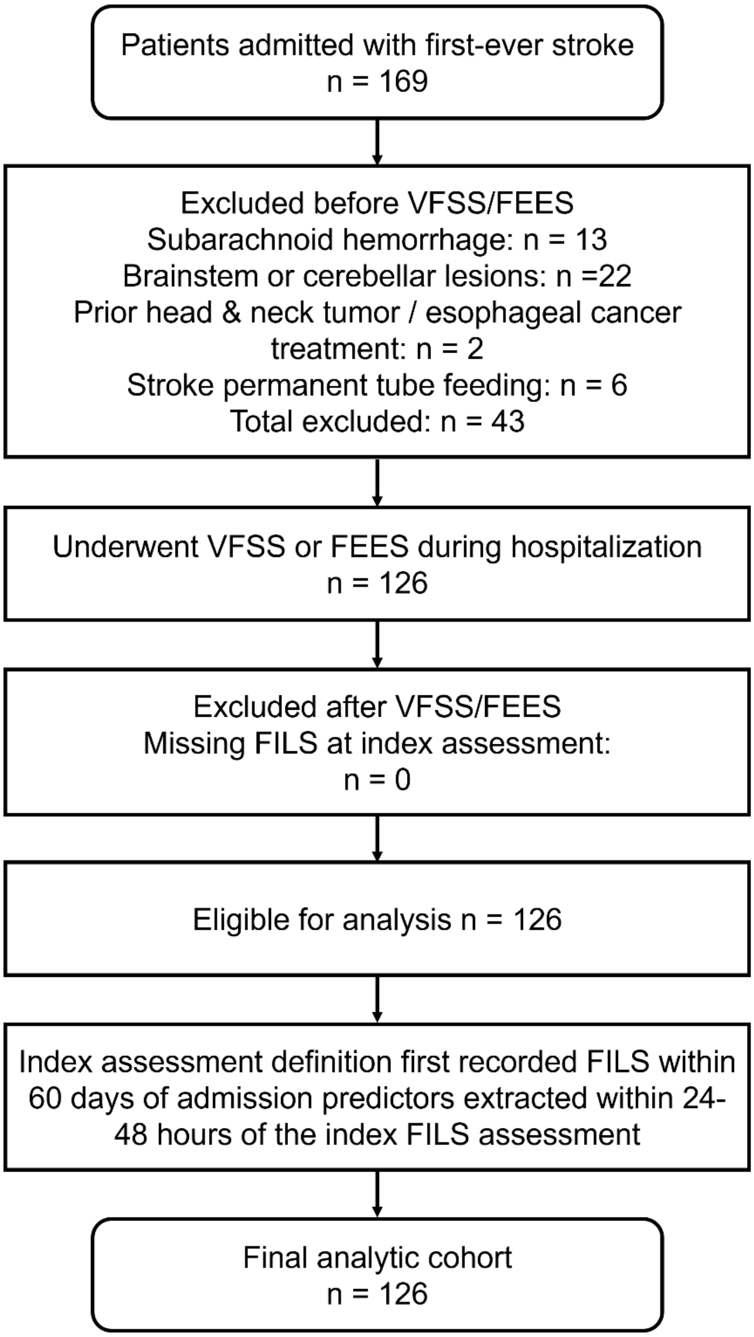


**Figure S1.** Flow diagram of patient identification, exclusions, and the final analytic cohort. FILS indicates the Food Intake LEVEL Scale; VFSS indicates videofluoroscopic swallow study; FEES indicates fiberoptic endoscopic evaluation of swallowing.

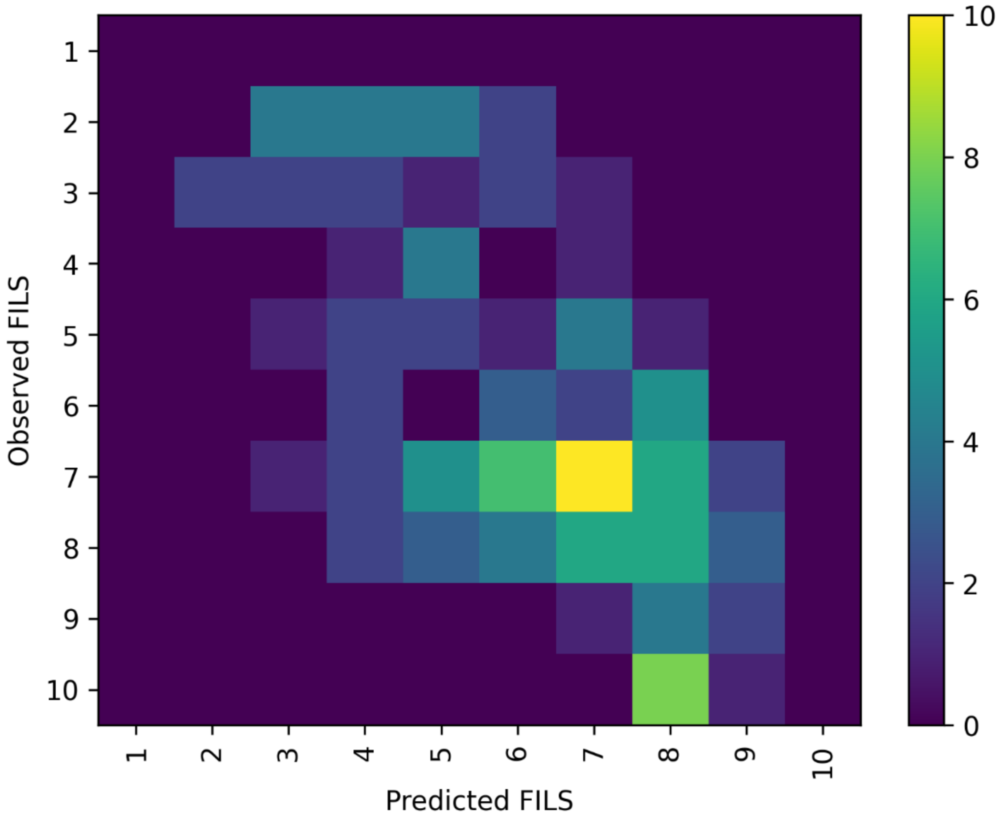


**Figure S2.** Confusion matrix (heatmap) for observed versus predicted the Food Intake LEVEL Scale (FILS) categories from the supplementary ordinal analysis. Predictions are out-of-fold estimates from internal validation. This analysis is supplementary to the primary threshold-based models.

**Table S1. Sensitivity analyses (nested cross-validation; out-of-fold predictions).**

| Outcome | Analysis | n | Dropped | AUC mean (SD) | Brier mean |
|---|---|---|---|---|---|
| FILS ≥3 | Primary | 126 | 0 | 0.777 (0.102) | 0.095 |
| | Complete-case | 126 | 0 | 0.777 (0.102) | 0.095 |
| | No class weighting | 126 | 0 | 0.772 (0.108) | 0.095 |
| FILS ≥7 | Primary | 126 | 0 | 0.833 (0.065) | 0.167 |
| | Complete-case | 126 | 0 | 0.833 (0.065) | 0.167 |
| | No class weighting | 126 | 0 | 0.833 (0.063) | 0.167 |

**Abbreviations:** FILS, the Food Intake LEVEL Scale; AUC, area under the receiver operating characteristic curve.

Complete-case analysis excludes observations with missing values in any prespecified predictor. No class weighting repeats model development without class weighting to assess the impact of imbalance handling on probabilistic outputs. AUC mean (SD) represents the average of fold-specific AUC values across the outer cross-validation folds; therefore, it may differ slightly from the pooled out-of-fold AUC reported in Table 2.
